# Detection of the Novel SARS-CoV-2 European Lineage B.1.177 in Ontario, Canada

**DOI:** 10.1101/2020.11.30.20241265

**Authors:** Jennifer L. Guthrie, Sarah Teatero, Sandra Zittermann, Yao Chen, Ashleigh Sullivan, Heather Rilkoff, Esha Joshi, Karthikeyan Sivaraman, Richard de Borja, Yogi Sundaravadanam, Michael Laszloffy, Lawrence Heisler, Vanessa G. Allen, Jared T. Simpson, Nahuel Fittipaldi

## Abstract

**Background:** Travel-related dissemination of SARS-CoV-2 continues to contribute to the global pandemic. A novel SARS-CoV-2 lineage (B.1.177) reportedly arose in Spain in the summer of 2020, with subsequent spread across Europe linked to travel by infected individuals. Surveillance and monitoring through the use of whole genome sequencing (WGS) offers insights into the global and local movement of pathogens such as SARS-CoV-2 and can detect introductions of novel variants.

**Methods:** We analyzed the genomes of SARS-CoV-2 sequenced for surveillance purposes from specimens received by Public Health Ontario (Sept 6 – Oct 10, 2020), collected from individuals in eastern Ontario. Taxonomic lineages were identified using pangolin (v2.08) and phylogenetic analysis incorporated publicly available genomes covering the same time period as the study sample. Epidemiological data collected from laboratory requisitions and standard reportable disease case investigation was integrated into the analysis.

**Results:** Genomic surveillance identified a COVID-19 case with SARS-CoV-2 lineage B.1.177 from an individual in eastern Ontario in late September, 2020. The individual had recently returned from Europe. Genomic analysis with publicly available data indicate the most closely related genomes to this specimen were from Southern Europe. Genomic surveillance did not identify further cases with this lineage.

**Conclusions:** Genomic surveillance allowed for early detection of a novel SARS-CoV-2 lineage in Ontario which was deemed to be travel related. This type of genomic-based surveillance is a key tool to measure the effectiveness of public health measures such as mandatory self-isolation for returned travellers, aimed at preventing onward transmission of newly introduced lineages of SARS-CoV-2.

## INTRODUCTION

Severe acute respiratory syndrome coronavirus 2 (SARS-CoV-2), the causative agent of coronavirus disease 2019 (COVID-19), emerged in China in late 2019 and quickly spread globally [1]. Since then, considerable efforts have been made to sequence SARS-CoV-2 genomes from clinical samples to monitor the virus as it moves within and across populations [2,3]. Due to proofreading capabilities during replication, SARS-CoV-2 accumulates mutations at a relatively slow estimated rate of 0.8 × 10^−3^ mutations per site-year [4]. These mutations enable classification into viral lineages and serve to identify emergence of novel variants. As of November 2020, there have been 4 major lineages and several hundred sublineages defined using the Phylogenetic Assignment of Named Global Outbreak LINeages (pangolin) algorithm [5]. A novel SARS-CoV-2 lineage (B.1.177), characterized by three single nucleotide polymorphisms (SNPs) (C22227T, C28932T, and G29645T), reportedly arose in Spain in the early summer of 2020, with subsequent spread across Europe linked to travel by infected individuals [6].

Travel between many countries has been restricted or banned in an attempt to control the spread of the pandemic. In response to COVID-19, a Canada-wide travel advisory was announced on March 14, 2020 recommending against non-essential travel outside Canada, and limiting the number of airports with international flights. Returning travelers are required to self-isolate for 14-days and seek testing where necessary [7]. Here, we report introduction of the B.1.177 lineage into Ontario, Canada by a traveller returned from Europe in late September 2020. Public health measures enabled rapid identification and apparent restriction of dissemination of this new SARS-CoV-2 variant.

## MATERIALS AND METHODS

### Study sample and data collection

Public Health Ontario (PHO) conducts SARS-CoV-2 diagnostic testing as part of an Ontario COVID-19 testing laboratory network. Specimens are received at PHO from multiple healthcare settings, including COVID-19 assessment centres, clinics, and hospitals located across the province. Demographic information, travel history and details of the specimen type and date of collection are captured on test requisitions at the time of specimen collection. Epidemiological data from standard reportable disease case investigation were extracted from Ontario’s integrated Public Health Information System (iPHIS). The study sample consisted of 111 SARS-CoV-2 specimens with genomes sequenced for surveillance purposes and received by PHO (September 6 – October 10, 2020), collected from individuals residing in four health administrative units in eastern Ontario. This represents 3.9% of all confirmed positive specimens in the region during this time period.

### Whole genome sequencing and bioinformatics analysis

RNA was extracted from SARS-CoV-2 positive specimens and cDNA was synthesized, following standard methods. Next, ARTIC V3 amplicons were generated and genomic libraries prepared using the Nextera XT DNA Library Preparation Kit (Illumina, San Diego, CA) [8,9]. Genomes were sequenced as paired-end (2×150-bp) reads on an Illumina MiSeq instrument. Bioinformatic analysis used a modified version of the COVID-19 Genomics UK (COG-UK) consortium pipeline for reference-based (GenBank accession MN908947.3) assembly and variant calling [10]. Sequencing quality control was performed using ncov-tools [11]. Consensus FASTA sequences with ≤1% ambiguous bases were aligned with MAFFT (v7.471)[12], 5’ and 3’ ends (positions 1–54 and 29837–29903 in the reference, respectively) were masked. Maximum-likelihood phylogenetic trees were constructed using IQ-TREE (v2.0.3) with –fconst used to specify the number of invariant sites [13]. SARS-CoV-2 lineages were categorized with pangolin v2.08 (pangoLEARN 2020-10-30) [14]. Consensus sequences were uploaded to GISAID (https://www.gisaid.org/) and accession numbers are available in **Table S1**. A random sample from complete genomes (>29,000bp) with high-coverage (<1% Ns) of the most common SARS-CoV-2 B.1 descendant lineage genomes with specimen collection dates from September 6 through October 10, 2020 were downloaded from GISAID on November 9, 2020. Alignment, masking and phylogenetic analysis were conducted as described above for PHO-generated genomes.

### ETHICS STATEMENT

PHO Ethics Review Board has determined that this project is exempt from research ethics committee review, as it describes analyses that were completed at PHO Laboratory as part of routine clinical respiratory testing during the COVID-19 pandemic in Ontario and are therefore considered public health practice.

## RESULTS

Analysis of SARS-CoV-2 genome sequences generated for provincial surveillance purposes identified an individual infected with a SARS-CoV-2 lineage B.1.177 in eastern Ontario. Review of records revealed that this individual had recently returned to Canada from travel to Europe. Symptoms typical of COVID-19 such as fever, sore throat and fatigue were reported to have started during the travel period. A nasopharyngeal specimen was collected at a COVID-19 assessment centre (in-person, ambulatory care) a few days following return travel to Canada. The specimen tested positive for SARS-CoV-2 with an rRT-PCR cycle threshold of 23.7. WGS analysis identified that this genome belonged to the recently emerged European B.1.177 lineage characterized by the presence of 3 SNPs (C22227T, C28932T, G29645T). Phylogenetic analysis of an additional 110 SARS-CoV-2 genomes included in the WGS surveillance of eastern Ontario in the two-week period before and after the specimen (WGS Id: ON-PHL-20-02301) was collected, revealed that this genome sequence was unique and unrelated to any others circulating in the region (i.e., lineages B.1.1, B.1.1.10, B.1.1.32, B.1.2, B.1.3, **Figure 1**). Samples had, on average, 99.6% genome coverage and 0–35 SNPs (mean 12.1, SD±7.4) between any pair of genomes. Notably, we did not identify onward transmission of this lineage in the following weeks within the region, which is compatible with epidemiological data suggesting the individual adhered to quarantine regulations. Comparing the genome of ON-PHL-20-02301 to publicly available global SARS-CoV-2 sequences collected over the study period demonstrated that this genome clustered with B.1.177 lineage sequences (**Figure 2**). The most closely related genomes were 4 SNPs different relative to ON-PHL-20-02301, and were collected from individuals in Southern Europe.

**Figure 1.**
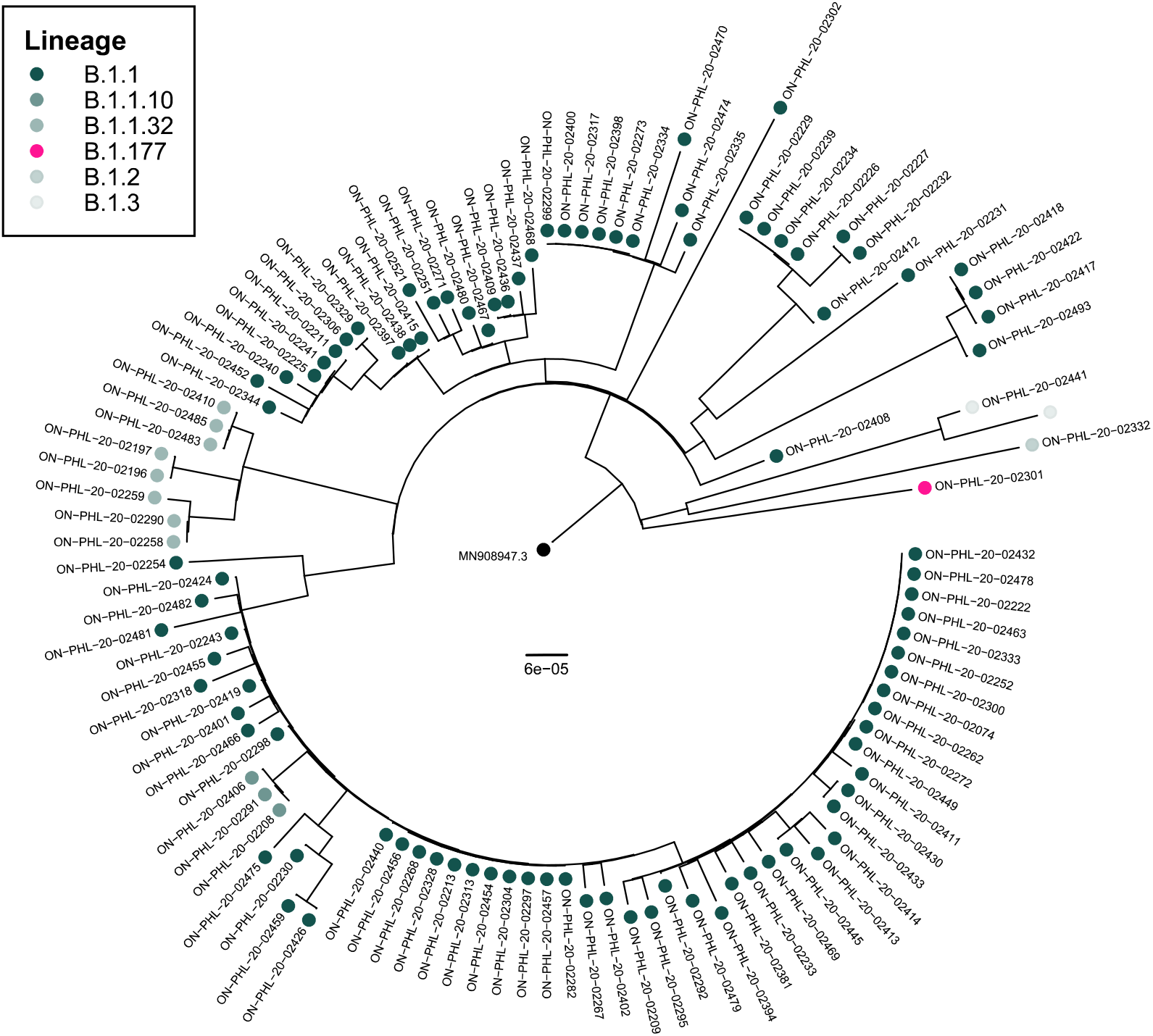
Phylogenetic analysis of SARS-CoV-2 genomes from individuals residing in four health administrative units in eastern Ontario, Canada, diagnosed with COVID-19 and submitted to Public Health Ontario from September 6, 2020 through October 10, 2020 (*n*=111). Tip points are coloured by lineage (https://github.com/cov-lineages/pangolin)

**Figure 2.**
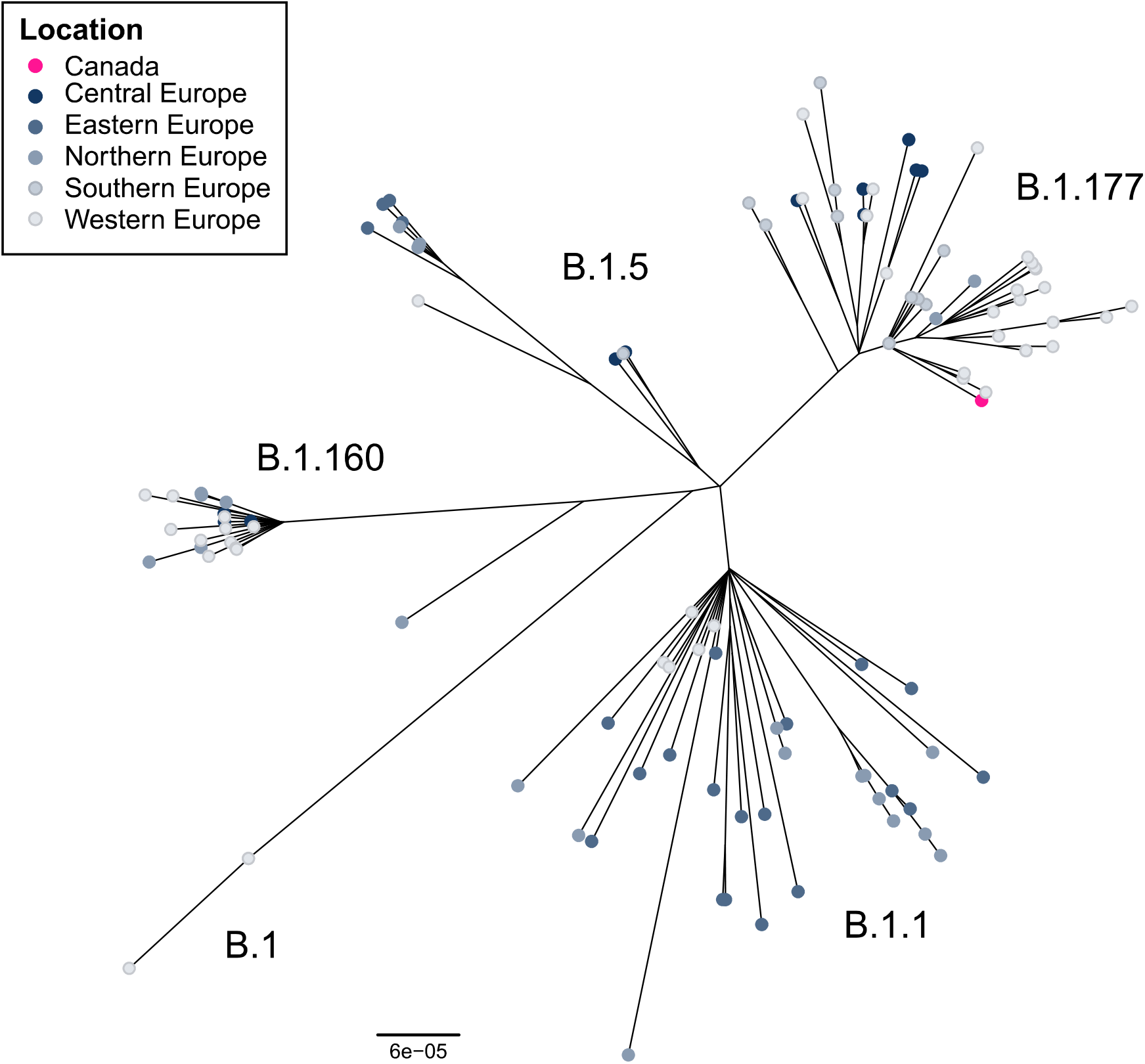
Unrooted phylogenetic tree of SARS-CoV-2 genome sequences, randomly selected from the most common B.1 descendant lineages with specimen collection dates of September 6, 2020 through October 10, 2020 (*n*=120) and deposited in GISAID (https://www.gisaid.org/) as of November 9, 2020. Included in the tree is the lineage B.1.177 SARS-CoV-2 genome from a specimen collected in Ontario, Canada. Tip points are coloured by location of specimen collection.

## DISCUSSION

Travellers contribute significantly to the spread of pathogens globally [15]. Recently, a newly described SARS-CoV-2 lineage B.1.177 emerged and disseminated across Europe during the summer of 2020. Here, we report the first described introduction of lineage B.1.177 into Ontario, Canada. Investigation revealed the individual had travelled to Europe and genomic epidemiological evidence suggests they likely acquired the infection abroad.

Despite travel restrictions in many countries, including Canada, international movement of persons has not been stopped in full, and dissemination of newly emerged viral variants is likely to continue. Effective prevention of the spread of novel SARS-CoV-2 variants requires strong public health measures, such as robust local testing capacity and WGS-based surveillance [3,16]. In this case, the combination of compliance with quarantine orders for returned travellers, accessibility of testing, and a provincial WGS-based surveillance system allowed for rapid identification and apparent prevention of dissemination of this variant in Ontario, thus supporting these public health measures. WGS analysis distinguished this genome from other regional cases during the same time period and identified viruses from Southern Europe as the closest genetic relatives. It should be noted that we cannot rule out onward transmission of this lineage as it is not possible to sequence every specimen due to availability of samples, and/or technical challenges related to low viral concentrations (high Ct specimens) [16]. However, a search of all GISAID submitted sequences did not reveal other B.1.177 genomes in Canada as of 30 November 2020.

Public health is under provincial and territorial jurisdiction in Canada, and epidemiological and clinical data sharing between different regions is challenging. Our findings highlight the important role of a provincial WGS-based surveillance program in COVID-19 response. It also makes a compelling case to further support a recently initiated national genomic surveillance effort (https://www.genomecanada.ca/en/cancogen), by removing barriers limiting the exchange of data between jurisdictions.

## Data Availability

Consensus genome sequences and metadata generated for and/or analysed as part of the study are available from GISAID (https://www.gisaid.org/).

## ACKNOWLEDGEMENTS

We acknowledge the contributions of the Virus Detection and Molecular Diagnostics sections of Public Health Ontario, and in particular Brenda Stanghini and Janet Folkes for specimen handling. We gratefully acknowledge contributions of SARS-CoV-2 genome sequences from other laboratories through GISAID (**Supplementary Table S2**).

