## Supplemental Tables for "Detection of the Novel SARS-CoV-2 European Lineage B.1.177 in Ontario, Canada"

### SUPPLEMENTARY MATERIALS

**Table S1.** Study sample sequence information.

| WGS Id | GISAID Accession | Specimen Collection | Lineage |
|---|---|---|---|
| ON-PHL-20-02074 | EPI_ISL_671605 | 2020-09 | B.1.1 |
| ON-PHL-20-02262 | EPI_ISL_671501 | 2020-09 | B.1.1 |
| ON-PHL-20-02196 | EPI_ISL_671496 | 2020-09 | B.1.1.32 |
| ON-PHL-20-02197 | EPI_ISL_671497 | 2020-09 | B.1.1.32 |
| ON-PHL-20-02258 | EPI_ISL_671498 | 2020-09 | B.1.1.32 |
| ON-PHL-20-02222 | EPI_ISL_671500 | 2020-09 | B.1.1 |
| ON-PHL-20-02259 | EPI_ISL_671499 | 2020-09 | B.1.1.32 |
| ON-PHL-20-02227 | EPI_ISL_671502 | 2020-09 | B.1.1 |
| ON-PHL-20-02234 | EPI_ISL_671506 | 2020-09 | B.1.1 |
| ON-PHL-20-02231 | EPI_ISL_671503 | 2020-09 | B.1.1 |
| ON-PHL-20-02233 | EPI_ISL_671504 | 2020-09 | B.1.1 |
| ON-PHL-20-02232 | EPI_ISL_671505 | 2020-09 | B.1.1 |
| ON-PHL-20-02241 | EPI_ISL_671512 | 2020-09 | B.1.1 |
| ON-PHL-20-02230 | EPI_ISL_671515 | 2020-09 | B.1.1 |
| ON-PHL-20-02243 | EPI_ISL_671513 | 2020-09 | B.1.1 |
| ON-PHL-20-02240 | EPI_ISL_671511 | 2020-09 | B.1.1 |
| ON-PHL-20-02213 | EPI_ISL_671509 | 2020-09 | B.1.1 |
| ON-PHL-20-02209 | EPI_ISL_671508 | 2020-09 | B.1.1 |
| ON-PHL-20-02211 | EPI_ISL_671507 | 2020-09 | B.1.1 |
| ON-PHL-20-02239 | EPI_ISL_671510 | 2020-09 | B.1.1 |
| ON-PHL-20-02229 | EPI_ISL_671514 | 2020-09 | B.1.1 |
| ON-PHL-20-02251 | EPI_ISL_671516 | 2020-09 | B.1.1 |
| ON-PHL-20-02208 | EPI_ISL_671520 | 2020-09 | B.1.1.10 |
| ON-PHL-20-02225 | EPI_ISL_671519 | 2020-09 | B.1.1 |
| ON-PHL-20-02254 | EPI_ISL_671518 | 2020-09 | B.1.1 |
| ON-PHL-20-02252 | EPI_ISL_671517 | 2020-09 | B.1.1 |
| ON-PHL-20-02226 | EPI_ISL_671595 | 2020-09 | B.1.1 |
| ON-PHL-20-02493 | EPI_ISL_671589 | 2020-09 | B.1.1 |
| ON-PHL-20-02313 | EPI_ISL_671551 | 2020-09 | B.1.1 |
| ON-PHL-20-02344 | EPI_ISL_671600 | 2020-09 | B.1.1 |
| ON-PHL-20-02306 | EPI_ISL_671550 | 2020-09 | B.1.1 |
| ON-PHL-20-02304 | EPI_ISL_671549 | 2020-09 | B.1.1 |
| ON-PHL-20-02318 | EPI_ISL_671553 | 2020-09 | B.1.1 |
| ON-PHL-20-02317 | EPI_ISL_671552 | 2020-09 | B.1.1 |
| ON-PHL-20-02329 | EPI_ISL_671555 | 2020-09 | B.1.1 |
| ON-PHL-20-02332 | EPI_ISL_671556 | 2020-09 | B.1.2 |
| ON-PHL-20-02333 | EPI_ISL_671601 | 2020-09 | B.1.1 |
| ON-PHL-20-02328 | EPI_ISL_671554 | 2020-09 | B.1.1 |
| ON-PHL-20-02272 | EPI_ISL_671560 | 2020-09 | B.1.1 |

| WGS Id | GISAID Accession | Specimen Collection | Lineage |
|---|---|---|---|
| ON-PHL-20-02335 | EPI_ISL_671567 | 2020-09 | B.1.1 |
| ON-PHL-20-02334 | EPI_ISL_671566 | 2020-09 | B.1.1 |
| ON-PHL-20-02302 | EPI_ISL_671565 | 2020-09 | B.1.1 |
| ON-PHL-20-02282 | EPI_ISL_671564 | 2020-09 | B.1.1 |
| ON-PHL-20-02268 | EPI_ISL_671563 | 2020-09 | B.1.1 |
| ON-PHL-20-02290 | EPI_ISL_671561 | 2020-09 | B.1.1.32 |
| ON-PHL-20-02271 | EPI_ISL_671559 | 2020-09 | B.1.1 |
| ON-PHL-20-02301 | EPI_ISL_671558 | 2020-09 | B.1.177 |
| ON-PHL-20-02300 | EPI_ISL_671557 | 2020-09 | B.1.1 |
| ON-PHL-20-02267 | EPI_ISL_671562 | 2020-09 | B.1.1 |
| ON-PHL-20-02292 | EPI_ISL_671522 | 2020-09 | B.1.1 |
| ON-PHL-20-02297 | EPI_ISL_671548 | 2020-09 | B.1.1 |
| ON-PHL-20-02273 | EPI_ISL_671547 | 2020-09 | B.1.1 |
| ON-PHL-20-02299 | EPI_ISL_671546 | 2020-09 | B.1.1 |
| ON-PHL-20-02295 | EPI_ISL_671523 | 2020-09 | B.1.1 |
| ON-PHL-20-02291 | EPI_ISL_671521 | 2020-09 | B.1.1.10 |
| ON-PHL-20-02298 | EPI_ISL_671524 | 2020-09 | B.1.1 |
| ON-PHL-20-02483 | EPI_ISL_671569 | 2020-09 | B.1.1.32 |
| ON-PHL-20-02485 | EPI_ISL_671568 | 2020-09 | B.1.1.32 |
| ON-PHL-20-02410 | EPI_ISL_671538 | 2020-09 | B.1.1.32 |
| ON-PHL-20-02455 | EPI_ISL_671570 | 2020-09 | B.1.1 |
| ON-PHL-20-02413 | EPI_ISL_671591 | 2020-09 | B.1.1 |
| ON-PHL-20-02408 | EPI_ISL_671590 | 2020-09 | B.1.1 |
| ON-PHL-20-02409 | EPI_ISL_671539 | 2020-09 | B.1.1 |
| ON-PHL-20-02456 | EPI_ISL_671599 | 2020-09 | B.1.1 |
| ON-PHL-20-02422 | EPI_ISL_671592 | 2020-09 | B.1.1 |
| ON-PHL-20-02424 | EPI_ISL_671594 | 2020-09 | B.1.1 |
| ON-PHL-20-02415 | EPI_ISL_671606 | 2020-09 | B.1.1 |
| ON-PHL-20-02454 | EPI_ISL_671545 | 2020-09 | B.1.1 |
| ON-PHL-20-02412 | EPI_ISL_671540 | 2020-09 | B.1.1 |
| ON-PHL-20-02457 | EPI_ISL_671598 | 2020-09 | B.1.1 |
| ON-PHL-20-02411 | EPI_ISL_671541 | 2020-09 | B.1.1 |
| ON-PHL-20-02445 | EPI_ISL_671544 | 2020-09 | B.1.1 |
| ON-PHL-20-02426 | EPI_ISL_671536 | 2020-09 | B.1.1 |
| ON-PHL-20-02430 | EPI_ISL_671593 | 2020-09 | B.1.1 |
| ON-PHL-20-02394 | EPI_ISL_671537 | 2020-09 | B.1.1 |
| ON-PHL-20-02446 | EPI_ISL_671543 | 2020-09 | B.1.3 |
| ON-PHL-20-02449 | EPI_ISL_671542 | 2020-09 | B.1.1 |
| ON-PHL-20-02406 | EPI_ISL_671596 | 2020-10 | B.1.1.10 |
| ON-PHL-20-02414 | EPI_ISL_671525 | 2020-10 | B.1.1 |
| ON-PHL-20-02397 | EPI_ISL_671526 | 2020-10 | B.1.1 |
| ON-PHL-20-02398 | EPI_ISL_671527 | 2020-10 | B.1.1 |
| ON-PHL-20-02418 | EPI_ISL_671528 | 2020-10 | B.1.1 |
| ON-PHL-20-02417 | EPI_ISL_671529 | 2020-10 | B.1.1 |

| WGS Id | GISAID Accession | Specimen Collection | Lineage |
|---|---|---|---|
| ON-PHL-20-02381 | EPI_ISL_671530 | 2020-10 | B.1.1 |
| ON-PHL-20-02419 | EPI_ISL_671531 | 2020-10 | B.1.1 |
| ON-PHL-20-02433 | EPI_ISL_671532 | 2020-10 | B.1.1 |
| ON-PHL-20-02400 | EPI_ISL_671533 | 2020-10 | B.1.1 |
| ON-PHL-20-02432 | EPI_ISL_671534 | 2020-10 | B.1.1 |
| ON-PHL-20-02452 | EPI_ISL_671602 | 2020-10 | B.1.1 |
| ON-PHL-20-02402 | EPI_ISL_671597 | 2020-10 | B.1.1 |
| ON-PHL-20-02466 | EPI_ISL_671576 | 2020-10 | B.1.1 |
| ON-PHL-20-02463 | EPI_ISL_671571 | 2020-10 | B.1.1 |
| ON-PHL-20-02467 | EPI_ISL_671575 | 2020-10 | B.1.1 |
| ON-PHL-20-02468 | EPI_ISL_671574 | 2020-10 | B.1.1 |
| ON-PHL-20-02401 | EPI_ISL_671535 | 2020-10 | B.1.1 |
| ON-PHL-20-02470 | EPI_ISL_671572 | 2020-10 | B.1.1 |
| ON-PHL-20-02469 | EPI_ISL_671573 | 2020-10 | B.1.1 |
| ON-PHL-20-02440 | EPI_ISL_671577 | 2020-10 | B.1.1 |
| ON-PHL-20-02441 | EPI_ISL_671578 | 2020-10 | B.1.3 |
| ON-PHL-20-02437 | EPI_ISL_671581 | 2020-10 | B.1.1 |
| ON-PHL-20-02436 | EPI_ISL_671580 | 2020-10 | B.1.1 |
| ON-PHL-20-02459 | EPI_ISL_671579 | 2020-10 | B.1.1 |
| ON-PHL-20-02438 | EPI_ISL_671582 | 2020-10 | B.1.1 |
| ON-PHL-20-02521 | EPI_ISL_671604 | 2020-10 | B.1.1 |
| ON-PHL-20-02474 | EPI_ISL_671588 | 2020-10 | B.1.1 |
| ON-PHL-20-02475 | EPI_ISL_671587 | 2020-10 | B.1.1 |
| ON-PHL-20-02478 | EPI_ISL_671586 | 2020-10 | B.1.1 |
| ON-PHL-20-02479 | EPI_ISL_671585 | 2020-10 | B.1.1 |
| ON-PHL-20-02481 | EPI_ISL_671584 | 2020-10 | B.1.1 |
| ON-PHL-20-02480 | EPI_ISL_671583 | 2020-10 | B.1.1 |
| ON-PHL-20-02482 | EPI_ISL_671603 | 2020-10 | B.1.1 |

**Table S2.** Sampled sequences of publicly available genomes from GISAID, downloaded November 9, 2020.

| GISAID Accession | Originating Laboratory | Submitting Laboratory | Authors |
|---|---|---|---|
| EPI_ISL_598557 | Lighthouse Lab in Milton Keynes | Wellcome Sanger Institute for the COVID-19 Genomics UK Consortium | Alderton et al. |
| EPI_ISL_582110, EPI_ISL_582116 | CNR Virus des Infections Respiratoires - France SUD | CNR Virus des Infections Respiratoires - France SUD | Bal et al. |
| EPI_ISL_613557 | CHRU Pontchaillou - Laboratoire de Virologie 2, rue Henri Le Guilloux | National Reference Center for Viruses of Respiratory Infections, Institut Pasteur, Paris | Barbet et al. |
| EPI_ISL_614282 | General practitioner | National Reference Center for Viruses of Respiratory Infections, Institut Pasteur, Paris | Barbet et al. |
| EPI_ISL_593908 | Labo Analyses Med, Sarcelles | National Reference Center for Viruses of Respiratory Infections, Institut Pasteur, Paris | Behillil et al. |
| EPI_ISL_593924 | Labo Analyses Med, Puteaux | National Reference Center for Viruses of Respiratory Infections, Institut Pasteur, Paris | Behillil et al. |
| EPI_ISL_593935 | Sentinelles, Plessis-Trevise | National Reference Center for Viruses of Respiratory Infections, Institut Pasteur, Paris | Behillil et al. |
| EPI_ISL_560433, EPI_ISL_560436, EPI_ISL_560441, EPI_ISL_560516,EPI_ISL_560519, EPI_ISL_560528, EPI_ISL_560547, EPI_ISL_560551, EPI_ISL_603314, EPI_ISL_603426, EPI_ISL_603427, EPI_ISL_603541 | Viollier AG | Department of Biosystems Science and Engineering, ETH Zürich | Beisel et al. |
| EPI_ISL_578263, EPI_ISL_578269, EPI_ISL_596898, EPI_ISL_596900, EPI_ISL_596901, EPI_ISL_596909, EPI_ISL_596910, EPI_ISL_596917, EPI_ISL_596920, EPI_ISL_596925, EPI_ISL_605076, EPI_ISL_605086, EPI_ISL_605102, EPI_ISL_605124 | National Virus Reference Laboratory | National Virus Reference Laboratory | Carr et al. |
| EPI_ISL_582052, EPI_ISL_582064, EPI_ISL_582066, EPI_ISL_582067, EPI_ISL_582068, EPI_ISL_582069, EPI_ISL_582075, EPI_ISL_582076, EPI_ISL_582078, EPI_ISL_582080, EPI_ISL_582081, EPI_ISL_582084, EPI_ISL_582090 | Servicio de Microbiología. Hospital Universitario Donostia. | SeqCOVID-SPAIN consortium/IBV(CSIC) | Cilla et al. |

| GISAID Accession | Originating Laboratory | Submitting Laboratory | Authors |
|---|---|---|---|
| EPI_ISL_615647, EPI_ISL_615664, EPI_ISL_615747, EPI_ISL_620829, EPI_ISL_620951, EPI_ISL_621045, EPI_ISL_621404, EPI_ISL_621429, EPI_ISL_621525, EPI_ISL_621583, EPI_ISL_621728, EPI_ISL_621776, EPI_ISL_622747 | Department of Virus and Microbiological Special Diagnostics, Statens Serum Institut, Denmark | Albertsen lab, Department of Chemistry and Bioscience, Aalborg University, Denmark | Danish Covid-19 Genome Consortia |
| EPI_ISL_584336 | Virology Department, Sheffield Teaching Hospitals NHS Foundation Trust/ | COVID-19 Genomics UK (COG-UK) Consortium | de Silva et al. |
| EPI_ISL_597512, EPI_ISL_608480 | Lighthouse Lab in Cambridge | Wellcome Sanger Institute for the COVID-19 Genomics UK Consortium | Howes et al. |
| EPI_ISL_602338, EPI_ISL_602346, EPI_ISL_602374, EPI_ISL_602376, EPI_ISL_602388, EPI_ISL_602397, EPI_ISL_602405, EPI_ISL_602443, EPI_ISL_602455, EPI_ISL_596263, EPI_ISL_596300, EPI_ISL_596358 | HELIX LLC | WHO National Influenza Centre Russian Federation | Komissarov et al |
| EPI_ISL_596244 | WHO National Influenza Centre Russian Federation | WHO National Influenza Centre Russian Federation | Komissarov, A. |
| EPI_ISL_596248 | WHO National Influenza Centre Russian Federation | WHO National Influenza Centre Russian Federation | Komissarov, A. |
| EPI_ISL_593893, EPI_ISL_593900 | CHU Purpan - Laboratoire de Virologie - Institut Fédératif de Biologie | CHU Purpan - Laboratoire de Virologie - Institut Fédératif de Biologie | Latour et al. |
| EPI_ISL_569220, EPI_ISL_569232 | MEPHI, Aix Marseille University | MEPHI, Aix Marseille University | Levaseur A. |
| EPI_ISL_577832, EPI_ISL_577834, EPI_ISL_577835, EPI_ISL_577865, EPI_ISL_577992, EPI_ISL_577994, EPI_ISL_578020, EPI_ISL_578034, EPI_ISL_578053, EPI_ISL_578059, EPI_ISL_578073 | Dutch COVID-19 response team | Erasmus Medical Center | Munnink et al. |
| EPI_ISL_626572, EPI_ISL_626574, EPI_ISL_626575, EPI_ISL_626576, EPI_ISL_626577, EPI_ISL_626581, EPI_ISL_626592, EPI_ISL_626595, EPI_ISL_626616 | The National Institute of Public Health | State Veterinary Institute Prague | Nagy et al. |

| GISAID Accession | Originating Laboratory | Submitting Laboratory | Authors |
|---|---|---|---|
| EPI_ISL_590884 | Foerde Hospital, Department of Microbiology | Norwegian Institute of Public Health, Department of Virology | Stene-Johansen et al. |
| EPI_ISL_590887 | Vestfold Hospital, Toensberg Department of Microbiology | Norwegian Institute of Public Health, Department of Virology | Stene-Johansen et al. |
| EPI_ISL_590891 | Akershus University Hospital, Department for Microbiology and Infectious Disease Control | Norwegian Institute of Public Health, Department of Virology | Stene-Johansen et al. |
| EPI_ISL_590964, EPI_ISL_590966 | Dept. of Medical Microbiology, Stavanger University Hospital, Helse Stavanger HF | Norwegian Institute of Public Health, Department of Virology | Stene-Johansen et al. |
| EPI_ISL_590972 | Dept. of Medical Microbiology, Stavanger University Hospital, Helse Stavanger HF | Norwegian Institute of Public Health, Department of Virology | Stene-Johansen et al. |
| EPI_ISL_590980 | Oslo University Hospital, Department of Medical Microbiology | Norwegian Institute of Public Health, Department of Virology | Stene-Johansen et al. |
| EPI_ISL_590990 | Ostfold Hospital Trust - Kalnes, Centre for Laboratory Medicine | Norwegian Institute of Public Health, Department of Virology | Stene-Johansen et al. |
| EPI_ISL_591010 | Unilabs Laboratory Medicine | Norwegian Institute of Public Health, Department of Virology | Stene-Johansen et al. |
| EPI_ISL_591012 | Department of Medical Microbiology - section Molde, Molde Hospital | Norwegian Institute of Public Health, Department of Virology | Stene-Johansen et al. |
| EPI_ISL_589232, EPI_ISL_598969, EPI_ISL_599218, EPI_ISL_599301, EPI_ISL_599440, EPI_ISL_599800 | Lighthouse Lab in Glasgow | Wellcome Sanger Institute for the COVID-19 Genomics UK Consortium | VanSteenhouse et al. |
| EPI_ISL_580088, EPI_ISL_580516, EPI_ISL_597759 | Lighthouse Lab in Alderley Park | Wellcome Sanger Institute for the COVID-19 Genomics UK Consortium | Wynn et al. |
